# Examining the associations between the food environment and dietary intake in British Columbia: A cross-sectional study

**DOI:** 10.1101/2025.02.22.25322725

**Authors:** Bill Zhao, Tamara R. Cohen, Jason Sutherland, Rafael Meza, Parveen Bhatti, Rachel A Murphy

## Abstract

This study aimed to characterize neighbourhood food environments in British Columbia (BC) and to determine if food environment characteristics were associated with fruits and vegetables (FV) intake. A cross-sectional study was conducted using geospatial food environment (Can-FED) and neighbourhood environment measures from the BC Generations Project, a cohort of males and females aged 35-69. Food environment measures (chain grocery stores, fast food outlets, convenience stores, and two relative measures of food outlets) were categorized according to density: class 0 (no presence) to class 4 (highest density). Multivariable mixed-effect models adjusted for sociodemographics and neighbourhood characteristics were constructed to assess the associations between food environments and FV intakes as a continuous variable and as <5 or ≥ 5 servings of FV/day. Approximately 50% of participants lived in neighbourhoods without chain grocery stores, fast food outlets or convenience stores within walking distance. An inverse association between neighbourhoods with the highest density of fast-food restaurants and FV consumption was observed (Odds ratio (OR) =0.89, 95% CI: 0.80, 0.98). Associations between chain grocery stores, convenience stores and FV consumption were attenuated with adjustment for neighbourhood characteristics: walkability, material and social deprivation. The findings suggest the presence of food deserts across urban areas in BC. Participants living in neighbourhoods with greater density of fast-food restaurants were less likely to consume ≥5 servings of FV per day. Further studies are needed to understand the lack of associations between other characteristics of the food environment and dietary intake.

## Background

Poor diet quality is a leading cause of death and disability in high income countries, including Canada (1). Most Canadians consume low quality diets that do not meet guidelines (2). There is little evidence of improvement in diet quality over the past decade (3), and there is widening of inequities in diet quality by household income and education (4).

In 2016, Health Canada introduced the Healthy Eating Strategy aiming to improve the diet quality of Canadians (5). A central part of the strategy is improving the food environment to ensure healthy choices are easy, accessible, and affordable. The food environment encompasses the number and location of food outlets, often conceptualized as ‘food deserts’ and ‘food swamps’. The term food desert describes a geographical area with poor access to healthy and affordable foods (6). *Food deserts* typically exist in socioeconomically disadvantaged urban areas which may contribute to socioeconomic gradients in diet and diet-related health outcomes, although food deserts may also be present in rural areas where long distances are needed to access healthy food (7). A *food swamp* is a geographical area which has adequate access to healthier, affordable food with an abundance of food outlets that sell less healthy food and beverages such as convenience stores or fast-food restaurants (7).

Despite the growing interest in the food environment, there are large gaps in evidence that make it unclear how to focus public health efforts. For instance, there is uncertainty about the existence of food deserts in Canada, and subsequently on the role of food deserts on dietary intake. A study in Montreal, Quebec noted a lack of food deserts (8). Subsequently, a literature review highlighted evidence of inequities in food access in the United States but concluded studies in other high-income countries (including Canada) were limited and inconclusive (9). Studies of the food environment in Canada have largely focused on a single city/small region limiting applicability to other areas, and many are dated (8,10–13), representing non-current retail food environments.

Neighbourhood-level factors encompass the population composition and the built environment such as roads, parks and buildings. A study across multiple countries, including Canada, found that living in socioeconomically advantaged neighborhoods was associated with greater FV intake (14). Low socioeconomic status (SES) neighborhoods typically have fewer healthy food outlets and more stores offering unhealthy foods (15). In addition, unhealthy foods are usually less expensive than healthy foods (15). The structural hypothesis posits that the physical surroundings such as institutions and social norms of a neighbourhood can influence residents’ health behaviours (16). Additionally, the stress theory suggests that low SES neighbourhoods create stressful environments that may contribute to adopting unhealthy behaviours (16). However, few studies have considered neighbourhood-level factors in the context of associations between the food environment and dietary intake (17,18). To meet these research gaps, this study aimed to 1) characterize the food environment of participants in a large cohort study in British Columbia, and 2) determine if characteristics of the food environment were associated with FV consumption.

## Methods

Ethics approval was granted by the University of British Columbia, BC Cancer Research Ethics Board (H23-02251). Access to the de-identified participant data was granted Sept 5, 2023. Informed consent was obtained from all participants. This cross-sectional study was nested within the British Columbia Generations Project (BCGP) (19). Between 2009 and 2016, n=29,850 adults aged 35-69 were recruited from across BC. Most participants were from the Greater Vancouver area (Vancouver, Richmond, Coquitlam, and Burnaby) and Vancouver Island (Victoria and Nanaimo). At baseline, participants completed a detailed questionnaire providing information on personal demographics (e.g., household income, biological sex, ethnicity), health history (e.g., pre-existing conditions), and health behaviours (e.g., alcohol consumption, physical activity level, FV intake, sleep). All participants were asked to self-report measures of height and weight on the baseline questionnaire. Approximately 50% of participants also visited assessment centers where height and weight were measured by trained study staff. The dependent variable for this study was usual daily FV consumption. Participants were asked ‘In a typical day, how many total servings of fruit (not including fruit juice) do you eat?’ and ‘In a typical day, how many total servings of vegetables do you eat?’. One serving was defined as 1/2 cup or 125 ml of fresh, frozen, canned or cooked leafy vegetables. The number of servings of fruits and vegetables was then summed and explored as continuous servings, and dichotomized as <5 servings or ≥5 servings, in line with public health recommendations.

### Food Environment

The Canadian Food Environment Dataset (Can-FED) provides a detailed, nationwide examination of the retail food environment in Canada, leveraging data from the Statistics Canada Business Register from 2018 (20). This dataset originated from mandatory tax data collected by the Canada Revenue Agency. Can-FED data calculated density measures for food environments at the level of dissemination areas (DAs); a small relatively stable geographic unit with an average population of 400 to 700 persons. Can-FED measures were calculated within a 1-kilometer around the DAs. We used the general-use Can-FED dataset focusing on three categorical absolute density measures and two categorical relative density measures. The absolute density measures included fast food stores, convenience stores, and chain grocery stores calculated by dividing the count of a given outlet by the area in square kilometers. The two relative density measures were developed by the Can-FED investigators-the modified Retail Food Environment Index (mRFEI) and the fast-food restaurant mix (Rmix). mRFEI reflects the ratio of stores that are more likely to sell nutritious food including FV (i.e. chain grocery stores and fruit and vegetable markets) relative to chain supermarkets, grocery stores, fruit and vegetable markets, fast-food outlets, convenience stores and convenience stores at a gas station (20). The classification of the accessibility of foods within stores aligns with evidence on shelf space of ‘healthy’ or ‘unhealthy’ foods in different types of food stores (21). Rmix reflects the proportion of fast-food outlets relative to the total of fast-food and full-service restaurants. Each variable was classified into ordinally-ranked categories: class 0, class 1, class 2, class 3, and class 4, with 0 indicating no access to the outlet type and class 4 representing the highest density. For absolute density measures (chain grocery store, fast food outlet, and convenience stores) classes 3 and 4 were combined due to low cell counts.

### Built environment

The Canadian Urban Environmental Health Research Consortium (CANUE) generates standardized, analysis-ready geospatial environmental measures for every postal code in Canada (22). Variables of interest in this study were the Material and Social deprivation index (MSDI), neighbourhood walkability, and gentrification from the year 2016. The MSDI, generated at the DA level, reflects material and social dimensions of deprivation within communities using Canadian census data (23,24). The MSDI has been used extensively to monitor social inequalities in health, and has demonstrated construct validity. Material deprivation was determined by the proportion of individuals lacking a high school diploma, the employment-to-population ratio, and the mean income in the area (23,24). Social deprivation reflects the percentage of people living alone, the incidence of marital separation, and the percentage of single-parent families (23,24). Both measures were scored from 1 to 100, with 1 indicating lowest deprivation. Walkability measures were calculated using buffers of 1 km in radius, centered on the midpoint of DAs. Measures were then summarized using z-scores. Gentrification, a measure of changes in demographic and economic landscapes in a neighbourhood, was assessed using the GENUINE tool (25). This tool captured shifts in income, housing occupancy, educational attainment, and demographic trends from 2006 to 2016 to identify gentrification among Canadian metropolitan areas. Participants in the BCGP were linked with CANUE and Can-FED using six-digit postal codes for their primary residence provided at baseline.

### Covariates

Age, sex, ethnicity, education, marital status, and behaviours such as physical activity, smoking status, and alcohol consumption, were identified as possible confounders based on prior studies and directed acyclic graphs. Confounders were categorized according to established thresholds such as BMI, or from prior studies in the BCGP and/or CanPath cohorts (17,18,26).

### Study sample

Of the 29,580 participants enrolled in BCGP, 29,570 completed the BCGP baseline questionnaires. Of the 29,570 participants, 1,179 were excluded due to missing postal code information. Five duplicates were also removed, leaving a total of 28,386 participants. Of the 28,386 participants, food environment data were missing for 2,792 participants leaving 25,594 participants with complete information on food environment for aim 1. For aim 2, 598 participants were missing FV intake data, leaving a total of 24,996 participants for analysis.

### Statistical analysis

If physical activity from the long form IPAQ was not available, data from the short form IPAQ was used. When physical measurements of height and weight were not available, self-reported height and weight were used to calculate BMI. Otherwise, missing data were imputed using the ‘Multivariate Imputation by Chained Equations (MICE)’ package in R using predictive mean matching, logistic regression, and proportional odds logistic regression. Age and sex were correlated with missing data and included as auxiliary variables. In total, 20 sets of imputations were performed, with 10 iterations each to reach convergence. Imputed data were pooled using Rubin’s rule.

Descriptive statistics and cross-tabulations of measures of the food environment were performed. Linear mixed-effects models and generalized logistic mixed-effects models were used to investigate continuous and dichotomous consumption of FV, with nesting at the forward sortation area included as a random intercept. Each food environment measure (was modelled as an independent variable in separate models. Covariates were incorporated into the model using a forward stepwise approach. At each step, a new variable was added, and the model’s Akaike Information Criterion (AIC) was compared to that of the previous model. If the new model with the additional covariate exhibited a lower AIC, it was retained. This process continued until all covariates of interest were assessed. Model assumptions were verified using residual plots. Model 1 included adjustments for sex, age, income, marital status, education level, ethnicity, physical activity level, alcohol consumption, smoking status, BMI and urban/rural residential area. Model 2 additionally adjusted for neighborhood variables: urban/rural residential area, walkability, material deprivation, and social deprivation, to understand the effect on risk estimates between the food environment and FV intake. All analysis were conducted in R.

## Results

On average, participants were 55.8 years of age. Most participants (69%) were female, and Caucasian (86.1%). Approximately 53.1% of the participants earned an annual income of ≥$75,000 and 42.9% had a bachelor’s degree or higher. Most participants (39.9%) had a BMI <25kg/m^2^, and a substantial proportion (96.3%) lived in urban areas (Table 1). Descriptive statistics of participants by category (<5 vs. ≥5) of FV consumption are shown in Supplementary Table 1.

**Table 1.**
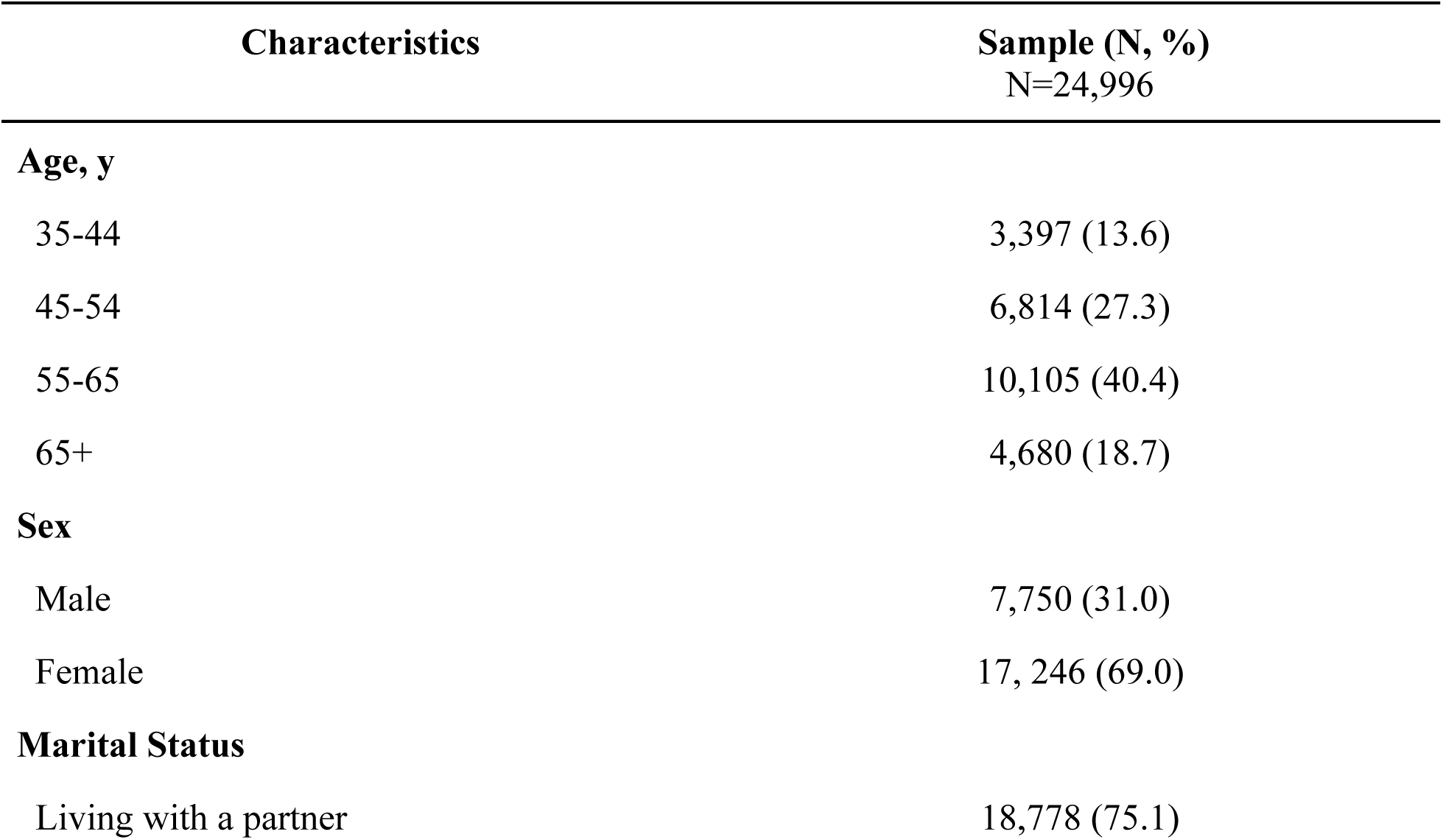
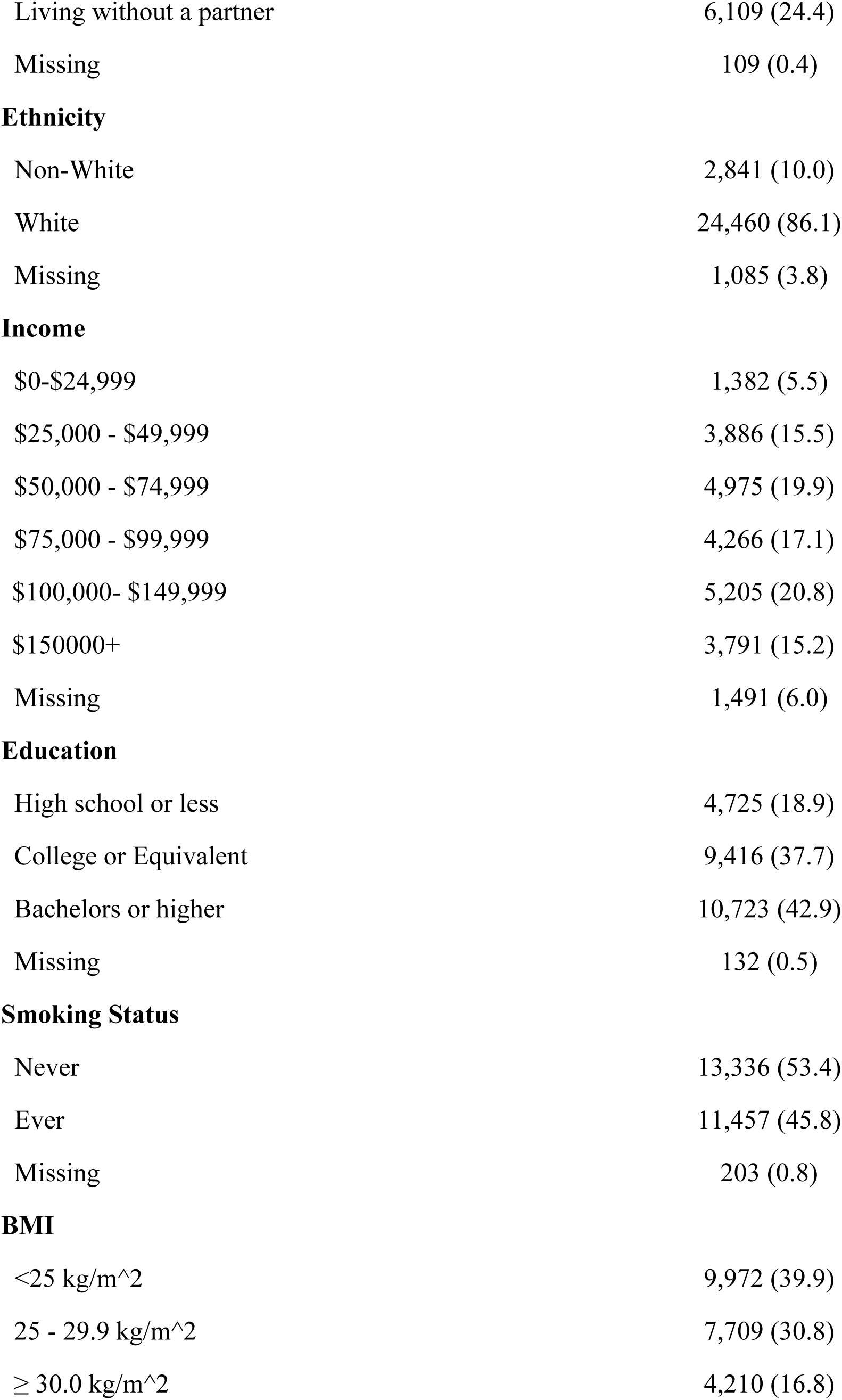
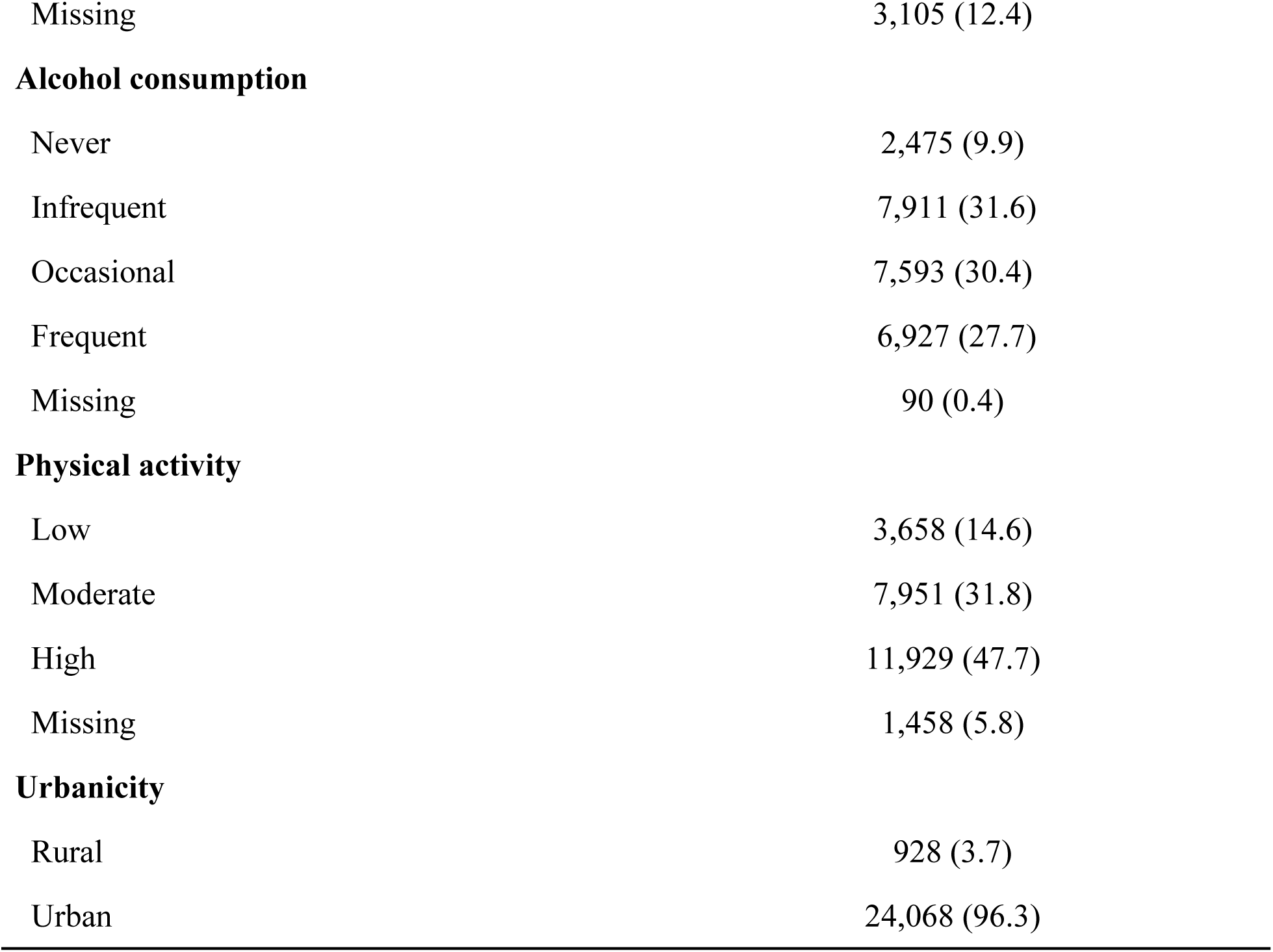
Characteristics of the BCGP participants in the analysis of the food environment and FV consumption.

| Characteristics | Sample (N, %)<br>N=24,996 |
| --- | --- |
| **Age, y** |  |
| 35-44 | 3,397 (13.6) |
| 45-54 | 6,814 (27.3) |
| 55-65 | 10,105 (40.4) |
| 65+ | 4,680 (18.7) |
| **Sex** |  |
| Male | 7,750 (31.0) |
| Female | 17, 246 (69.0) |
| **Marital Status** |  |
| Living with a partner | 18,778 (75.1) |
| Living without a partner | 6,109 (24.4) |
| Missing | 109 (0.4) |
| **Ethnicity** |  |
| Non-White | 2,841 (10.0) |
| White | 24,460 (86.1) |
| Missing | 1,085 (3.8) |
| **Income** |  |
| \$0-\$24,999 | 1,382 (5.5) |
| \$25,000 - \$49,999 | 3,886 (15.5) |
| \$50,000 - \$74,999 | 4,975 (19.9) |
| \$75,000 - \$99,999 | 4,266 (17.1) |
| \$100,000- \$149,999 | 5,205 (20.8) |
| \$150000+ | 3,791 (15.2) |
| Missing | 1,491 (6.0) |
| **Education** |  |
| High school or less | 4,725 (18.9) |
| College or Equivalent | 9,416 (37.7) |
| Bachelors or higher | 10,723 (42.9) |
| Missing | 132 (0.5) |
| **Smoking Status** |  |
| Never | 13,336 (53.4) |
| Ever | 11,457 (45.8) |
| Missing | 203 (0.8) |
| **BMI** |  |
| <25 kg/m<sup>2</sup> | 9,972 (39.9) |
| 25 - 29.9 kg/m<sup>2</sup> | 7,709 (30.8) |
| ≥ 30.0 kg/m<sup>2</sup> | 4,210 (16.8) |
| Missing | 3,105 (12.4) |
| **Alcohol consumption** |  |
| Never | 2,475 (9.9) |
| Infrequent | 7,911 (31.6) |
| Occasional | 7,593 (30.4) |
| Frequent | 6,927 (27.7) |
| Missing | 90 (0.4) |
| **Physical activity** |  |
| Low | 3,658 (14.6) |
| Moderate | 7,951 (31.8) |
| High | 11,929 (47.7) |
| Missing | 1,458 (5.8) |
| **Urbanicity** |  |
| Rural | 928 (3.7) |
| Urban | 24,068 (96.3) |

In general, a large portion of BCGP participants lacked access (class 0) to any retail food outlets: 48.3% had no access to outlets that sold FV (mRFEI), suggesting the presence of food deserts (Table 2). Over half of the participants (53.5%) had no fast-food restaurants where they lived, and 47.7% lacked access to convenience stores. In contrast, 11.6% of the participants lived in neighbourhoods with high densities of fast-food outlets (class 3/4), and 11.3% lived in neighbourhoods with high densities of convenience stores (class 3/4).

**Table 2.** The distribution of BCGP participants living in different food environments in BC.

| Food environment measure | Can-FED Classification** |
| --- | --- |
| N=25,594* | (N, %) |

|  | 0 | class 1 | class 2 | class 3 | class 4 |
| --- | --- | --- | --- | --- | --- |
| mRFEI | 12,360<br>(48.3) | 2,378<br>(9.3) | 3,863<br>(15.1) | 4,908<br>(19.2) | 2,085<br>(8.1) |
| Rmix | 13,698<br>(53.5) | 3,348<br>(13.1) | 2,524<br>(9.9) | 2,834<br>(11.1) | 3,190<br>(12.5) |
| Density of chain grocery stores | 19,393<br>(75.8) | 2,875<br>(11.2) | 1,502<br>(5.9) | 1,105<br>(4.3) | 719<br>(2.8) |
| Density of fast-food restaurants | 13,698<br>(53.5) | 5,363<br>(21.0) | 3,570<br>(13.9) | 2,447<br>(9.6) | 516<br>(2.0) |
| Density of convenience stores | 12,218<br>(47.7) | 6,236<br>(24.4) | 4,224<br>(16.5) | 2,521<br>(9.8) | 395<br>(1.5) |

Zero represents no access at all, Class 1 is the lowest density level and Class 4 is the highest density level. Density increases with increasing class.

Over half (50.8%) of participants had no immediate access to fast-food restaurants or chain grocery stores, while 4.5% lived in neighbourhoods with high densities of both (Table 3). Similarly, 44.1% of participants lived in neighbourhoods with no access to chain grocery stores or convenience stores, while 3.2% lived in neighbourhoods with high densities of both (Table 4).

**Table 3.**
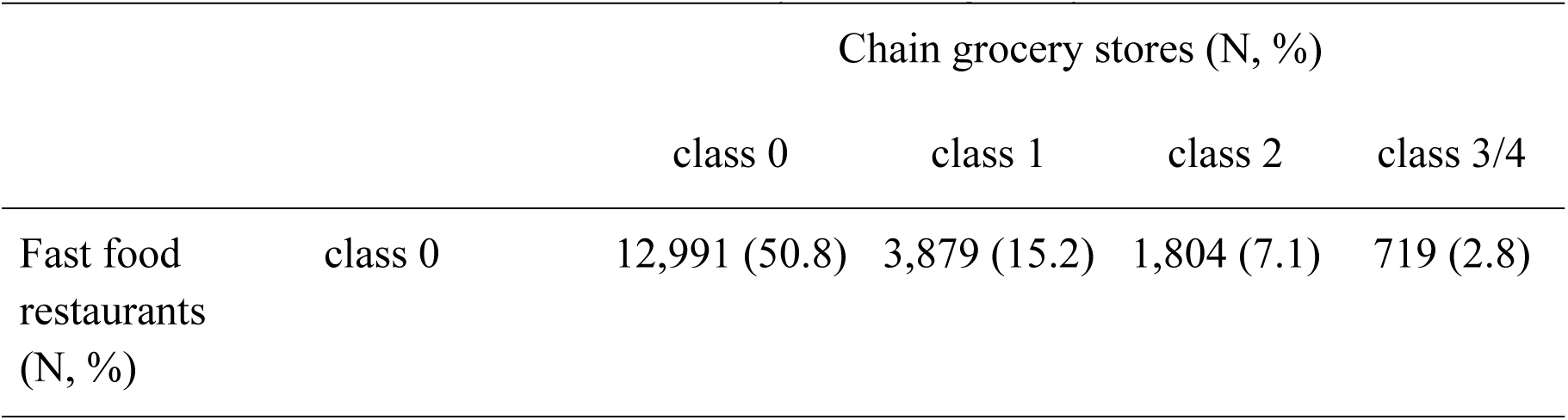
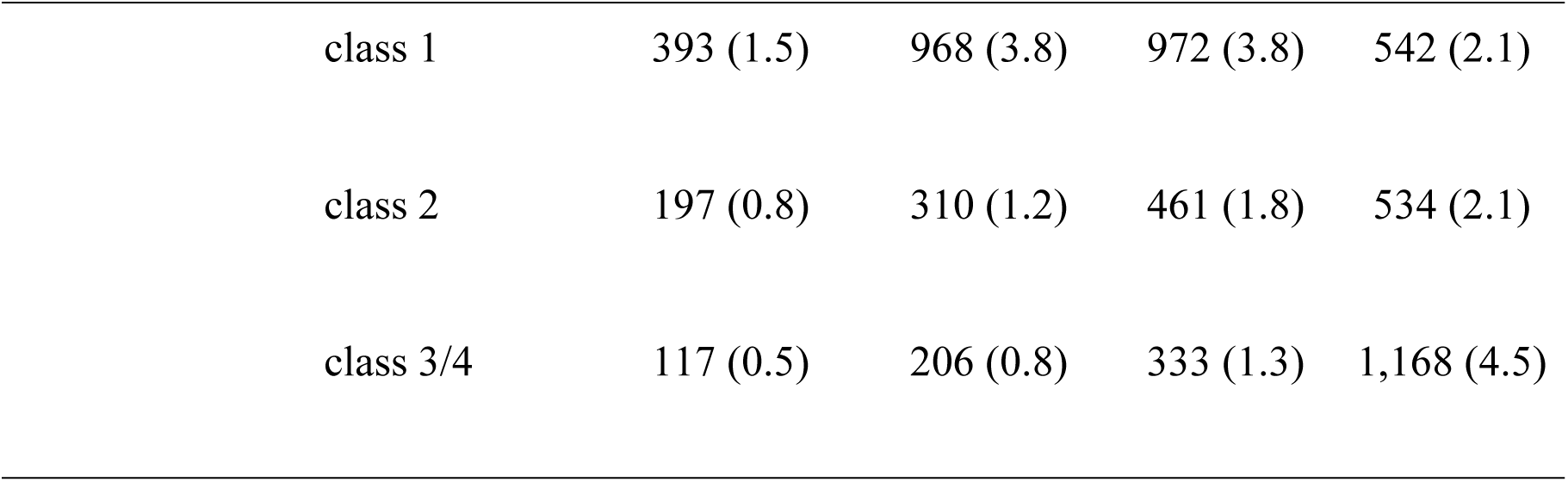
Cross-tabulation between the density of chain grocery stores and fast-food restaurants.

**Table 4.** Cross-tabulation between the density of chain grocery stores and convenience stores.

|  |  | Chain grocery stores (N, %) |  |  |  |
| --- | --- | --- | --- | --- | --- |
|  |  | class 0 | class 1 | class 2 | class 3/4 |
| Convenience stores<br>(N, %) | class 0 | 11,298 (44.1) | 4,355 (17.0) | 2,614 (10.2) | 1,126 (4.4) |
|  | class 1 | 411 (1.6) | 1021 (4.0) | 849 (3.3) | 603 (2.4) |
|  | class 2 | 243 (0.9) | 485 (1.9) | 406 (1.6) | 377 (1.5) |
|  | class 3/4 | 275 (1.1) | 384 (1.5) | 355 (1.4) | 810 (3.2) |

Only 6.1% of participants lived in areas with high densities of both fast-food outlets and convenience stores, while 39.6% of participants lived in neighbourhoods with no access to either (Table 5). Notably, neighbourhoods without access to chain grocery stores were not characterized by high density of fast food (0.5%) or convenience stores (1.1%).

**Table 5.** Cross-tabulation between the density of fast-food restaurants and convenience stores.

|  |  | class 0 | class 1 | class 2 | class 3/4 |
| --- | --- | --- | --- | --- | --- |
| Convenience stores<br>(N, %) | class 0 | 9,557 (37.3) | 2,607 (10.2) | 1,236 (4.8) | 298 (1.2) |
|  | class 1 | 1,638 (6.4) | 2,093 (8.2) | 1,225 (4.8) | 407 (1.6) |
|  | class 2 | 814 (3.2) | 1,024 (4.0) | 1,076 (4.2) | 656 (2.6) |
|  | class 3/4 | 209 (0.8) | 512 (2.0) | 687 (2.7) | 1,555 (6.1) |

The mean daily FV consumption for participants in BCGP was 5.3 servings. Males consumed a mean of 4.5 servings, while females consumed a mean of 5.7 servings. There were no significant differences in mean FV consumption across given food environment measures (Supplementary Table 2). The linear analysis did not reveal any associations between the food environment measures and consumption of FV (Table 6).

**Table 6.**
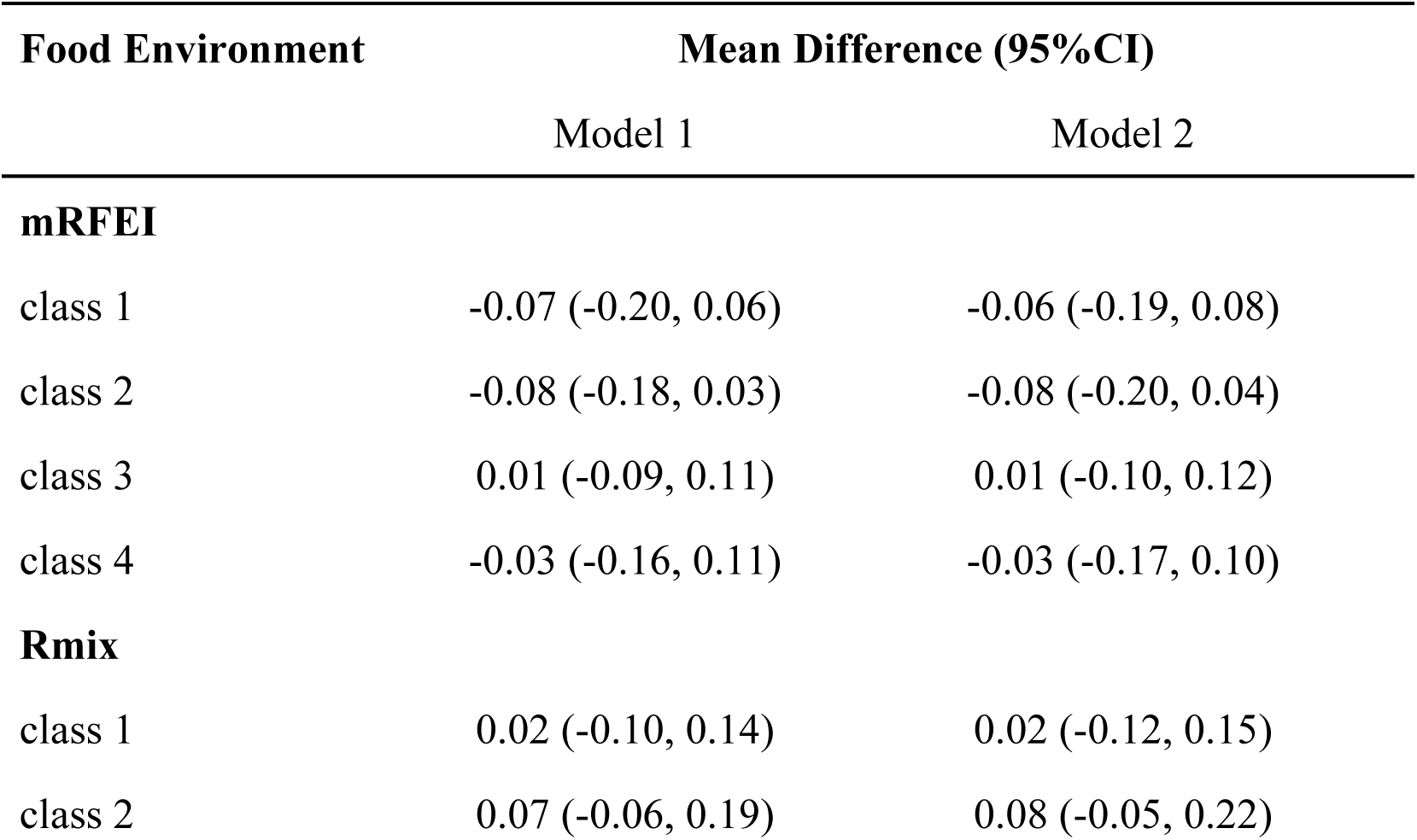
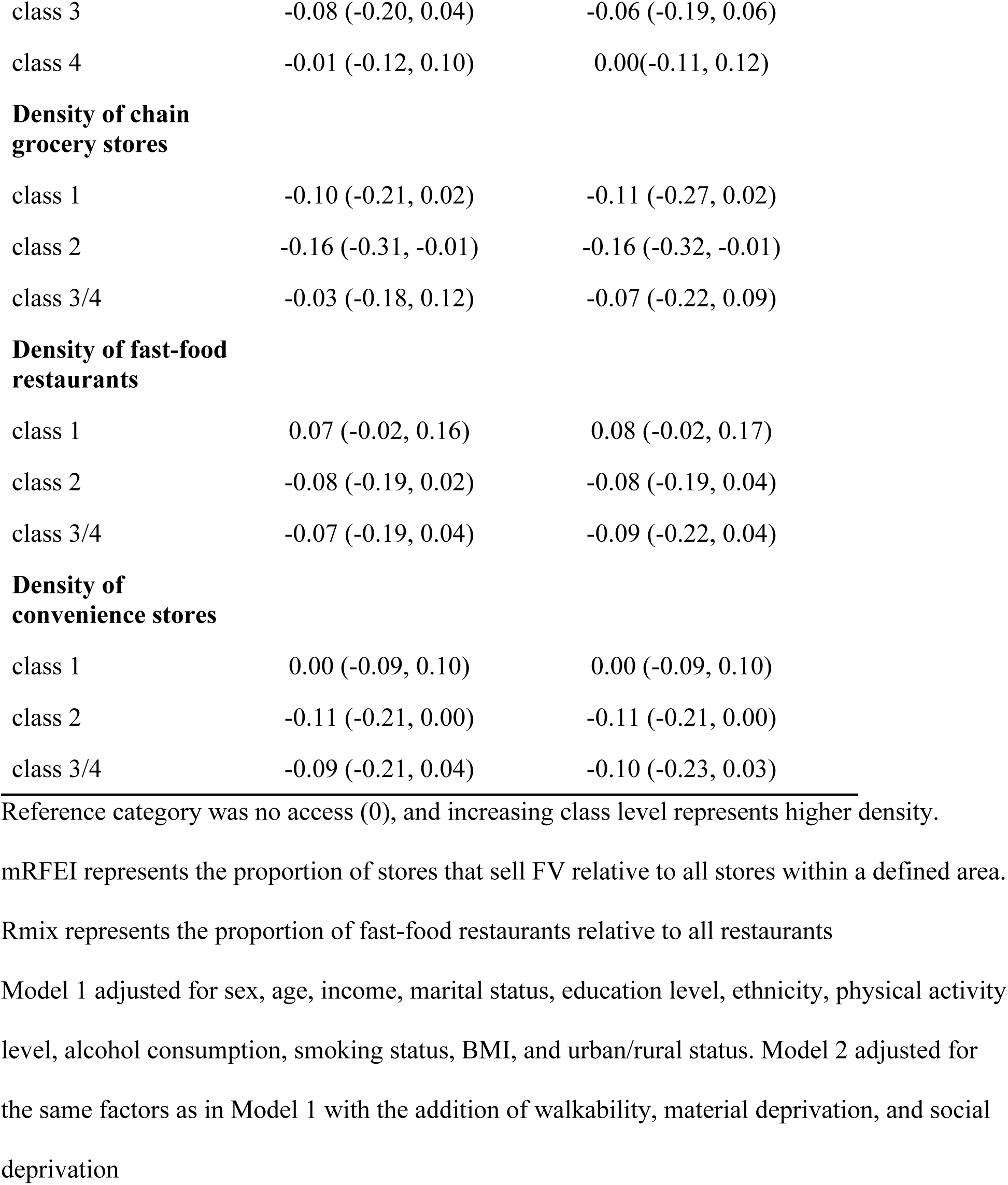
Mean difference in FV consumption (95% CI) for BCGP participants, N=24,996.

| Food Environment | Mean Difference (95%CI) |  |
| --- | --- | --- |
|  | Model 1 | Model 2 |
| **mRFEI** |  |  |
| class 1 | -0.07 (-0.20, 0.06) | -0.06 (-0.19, 0.08) |
| class 2 | -0.08 (-0.18, 0.03) | -0.08 (-0.20, 0.04) |
| class 3 | 0.01 (-0.09, 0.11) | 0.01 (-0.10, 0.12) |
| class 4 | -0.03 (-0.16, 0.11) | -0.03 (-0.17, 0.10) |
| **Rmix** |  |  |
| class 1 | 0.02 (-0.10, 0.14) | 0.02 (-0.12, 0.15) |
| class 2 | 0.07 (-0.06, 0.19) | 0.08 (-0.05, 0.22) |
| class 3 | -0.08 (-0.20, 0.04) | -0.06 (-0.19, 0.06) |
| class 4 | -0.01 (-0.12, 0.10) | 0.00(-0.11, 0.12) |
| **Density of chain grocery stores** |  |  |
| class 1 | -0.10 (-0.21, 0.02) | -0.11 (-0.27, 0.02) |
| class 2 | -0.16 (-0.31, -0.01) | -0.16 (-0.32, -0.01) |
| class 3/4 | -0.03 (-0.18, 0.12) | -0.07 (-0.22, 0.09) |
| **Density of fast-food restaurants** |  |  |
| class 1 | 0.07 (-0.02, 0.16) | 0.08 (-0.02, 0.17) |
| class 2 | -0.08 (-0.19, 0.02) | -0.08 (-0.19, 0.04) |
| class 3/4 | -0.07 (-0.19, 0.04) | -0.09 (-0.22, 0.04) |
| **Density of convenience stores** |  |  |
| class 1 | 0.00 (-0.09, 0.10) | 0.00 (-0.09, 0.10) |
| class 2 | -0.11 (-0.21, 0.00) | -0.11 (-0.21, 0.00) |
| class 3/4 | -0.09 (-0.21, 0.04) | -0.10 (-0.23, 0.03) |

Reference category was no access (0), and increasing class level represents higher density.
mRFEI represents the proportion of stores that sell FV relative to all stores within a defined area.
Rmix represents the proportion of fast-food restaurants relative to all restaurants
Model 1 adjusted for sex, age, income, marital status, education level, ethnicity, physical activity
level, alcohol consumption, smoking status, BMI, and urban/rural status. Model 2 adjusted for
the same factors as in Model 1 with the addition of walkability, material deprivation, and social
deprivation

In model 1 of the logistic analysis, participants living in areas with intermediate convenience store density (class 2) had lower odds of meeting recommendations for FV intake (Table 7). With adjustment for walkability, material and social deprivation in model 2, statistical significance was attenuated (OR 0.93, 95% CI 0.86, 1.01). Similarly, in model 1, participants living in neighbourhoods with intermediate mRFEI had lower odds of meeting recommendations for FV intake, but statistical significance was attenuated in model 2 (OR=0.94, 95% CI 0.86, 1.03). Participants living in neighbourhoods with intermediate chain grocery store density (class 2) had lower odds of consuming ≥5 servings of FV per day across all models. Generally, participants living in neighbourhoods with increasing density of fast-food restaurants had lower odds of meeting recommendations for FV. For instance, class 3/4 had an OR=0.89, 95% CI 0.80, 0.98 in model 2.

**Table 7.**
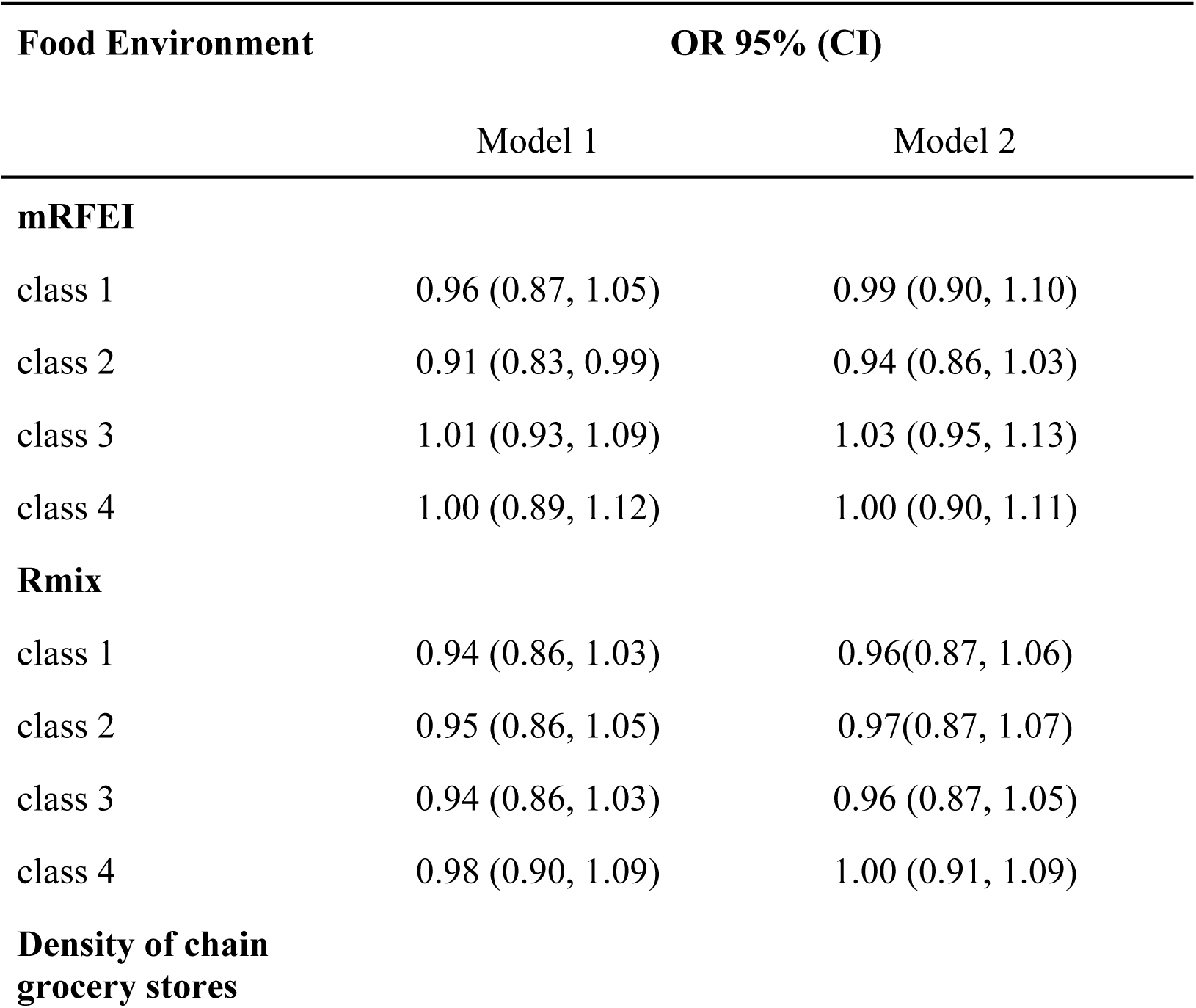
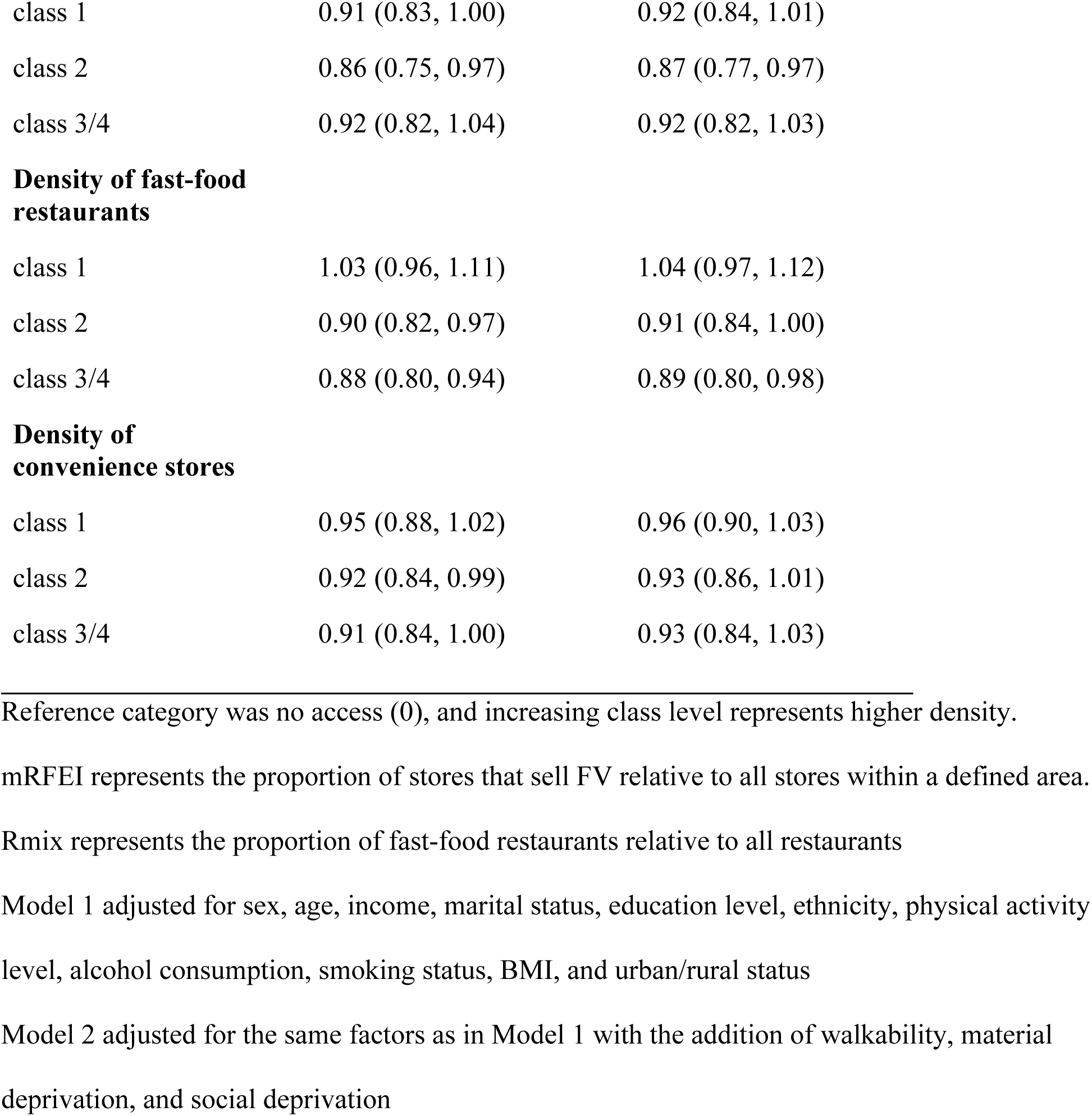
Odds ratios (OR, 95% CI) for consuming 5 servings/day or greater of FV for different food environment measures among BCGP participants, N=24,996.

| Food Environment | OR 95% (CI) |  |
| --- | --- | --- |
|  | Model 1 | Model 2 |
| **mRFEI** |  |  |
| class 1 | 0.96 (0.87, 1.05) | 0.99 (0.90, 1.10) |
| class 2 | 0.91 (0.83, 0.99) | 0.94 (0.86, 1.03) |
| class 3 | 1.01 (0.93, 1.09) | 1.03 (0.95, 1.13) |
| class 4 | 1.00 (0.89, 1.12) | 1.00 (0.90, 1.11) |
| **Rmix** |  |  |
| class 1 | 0.94 (0.86, 1.03) | 0.96(0.87, 1.06) |
| class 2 | 0.95 (0.86, 1.05) | 0.97(0.87, 1.07) |
| class 3 | 0.94 (0.86, 1.03) | 0.96 (0.87, 1.05) |
| class 4 | 0.98 (0.90, 1.09) | 1.00 (0.91, 1.09) |
| **Density of chain grocery stores** |  |  |
| class 1 | 0.91 (0.83, 1.00) | 0.92 (0.84, 1.01) |
| class 2 | 0.86 (0.75, 0.97) | 0.87 (0.77, 0.97) |
| class 3/4 | 0.92 (0.82, 1.04) | 0.92 (0.82, 1.03) |
| **Density of fast-food restaurants** |  |  |
| class 1 | 1.03 (0.96, 1.11) | 1.04 (0.97, 1.12) |
| class 2 | 0.90 (0.82, 0.97) | 0.91 (0.84, 1.00) |
| class 3/4 | 0.88 (0.80, 0.94) | 0.89 (0.80, 0.98) |
| **Density of convenience stores** |  |  |
| class 1 | 0.95 (0.88, 1.02) | 0.96 (0.90, 1.03) |
| class 2 | 0.92 (0.84, 0.99) | 0.93 (0.86, 1.01) |
| class 3/4 | 0.91 (0.84, 1.00) | 0.93 (0.84, 1.03) |

- Reference category was no access (0), and increasing class level represents higher density. mRFEI represents the proportion of stores that sell FV relative to all stores within a defined area. Rmix represents the proportion of fast-food restaurants relative to all restaurants Model 1 adjusted for sex, age, income, marital status, education level, ethnicity, physical activity level, alcohol consumption, smoking status, BMI, and urban/rural status Model 2 adjusted for the same factors as in Model 1 with the addition of walkability, material deprivation, and social deprivation

## Discussion

Our findings suggest the presence of food deserts across many populous areas of BC. Within this population of nearly 25,000 adults, a large proportion (48.3%) did not have access to any chain supermarkets, grocery stores or fruits and vegetable markets (mRFEI) within 10-15 minute walk and 75.8% lacked access to a chain grocery store in their neighbourhood. Although our study population was generally affluent, the geographic areas studied included neighbourhoods with high material and social deprivation, and thus the implications of food deserts may be broader than findings in our study population indicate. There was also evidence of food environments that may be more favourable for healthier dietary intake, with nearly 40% of participants living in neighbourhoods with no convenience stores or fast-food outlets. The prevalence of areas without access to a grocery but high access to fast-food outlets and convenience stores was minimal.

Access to chain grocery stores relative to other stores (mRFEI) and chain grocery stores were not significantly associated with FV consumption. Literature on access to grocery stores and FV intake are mixed. For instance, Pessoa et al. found that higher FV intake was associated with living in neighbourhoods with a higher density of healthy food outlets (27). However, the study by Pessoa et al. was conducted in Brazil, where the types of food outlets and food systems differ from those in Canada. Furthermore, the definition of “healthy food outlets” encompassed various types of outlets, including “open-air markets” and “restaurants with nutritious food,” which differed from the measures used in this study (27). A study in the US reported that in urban areas, access to supermarkets was positively associated with FV consumption (28), although access was measured as proximity to grocery stores rather than density as in our study.

A study in Quebec found no associations between grocery store proximity or density with FV consumption. Studies from Hong Kong (29) and the UK (30) also found no significant association between grocery store density with FV consumption. The lack of associations found in this study may reflect that physical proximity to grocery stores is just one of many factors that affect FV consumption. For instance, Aggarwal et al reported that participants travelled further distances to shop at stores outside of their neighbourhood environment if the cost of food was lower (31). Similarly, other studies found that food prices may be more associated with FV consumption than physical proximity (32). Studies from the US further suggested that increasing geographic access to FV by adding grocery stores in food deserts did not result in changes in FV consumption for residents (32).

Our findings suggest that participants living in neighbourhoods with greater fast-food outlets were less likely to meet FV recommendations, but the difference in FV consumption was minimal. Other studies have found that both high and intermediate density of fast-food outlets may be negatively associated with FV. A study in Hong Kong found that the highest quartile of fast-food density was associated with lower fruit consumption and a trend for decreasing fruit consumption with increasing density of fast-food outlets (29).

Given that convenience stores seldom sell FV (21), we expected convenience stores may influence dietary intake through greater access to energy dense, nutrient poor foods which might displace FV in the diet. However, statistically significant associations between convenience store density measures and FV consumption were attenuated with adjustment for neighborhood characteristics. In contrast, a study in New Zealand found that individuals living in areas with the shortest travel time to convenience stores had lower FV consumption (33). A higher proportion of fast-food restaurants and convenience stores relative to grocery stores was associated with lower FV consumption in a population of Hong Kong (29). A study in Quebec reported that people living in neighbourhoods with the highest density of convenience stores had greater FV consumption (34), although they attributed the finding to residual confounding by individual-level variables, particularly sex, income and age.

The use of data from the BCGP which spans major urban cities and rural regions in BC is a strength and fills a gap in evidence on the retail food environment in the most populous areas in BC. Previous studies conducted in select cities in Canada and the United States—where varying demographics, bylaws, and possibly unique food environments limits the applicability of findings to BC. Although the data was collected some ten years prior, this pre-dated the rise in online grocery delivery and food delivery service (35) that may blur boundaries of the food environment. Our results may serve as a ‘baseline’ to understand the effect of changes in food purchasing behaviours on food environment and dietary intake. Another strength of this study is the use of Can-FED, which provides comprehensive and standardized metrics of geographic access to different types of food outlets. A significant challenge in food environment research is the variability and inconsistent quality of the measurement of the food environment. For example, some studies measured public transportation and walking distance to stores (11) while others assessed distance using pathways over straight-line distances (36) or population-weighted distances (36). Studies also measured the number and type of food outlets in different manners such as through phone book listings (37) or commercial databases (10). These differing methods limit comparability across studies. Can-FED is a substantial step forward in addressing these issues by providing standardized nationwide measures. Can-FED also enables a more nuanced exploration of the food environment through the use of multiple measures beyond grocery stores and fast-food restaurants which predominate the literature (7).

This study also has limitations. The food environment was defined using DA boundaries based on the postal code of residence. However, people may perceive their neighbourhood differently and spend time and purchase food outside their residential neighbourhood (e.g., at work and leisure). It is thus possible the associations we observed were attenuated towards the null without consideration of perceived food environments and food environments beyond participants’ homes. The majority of BCGP participants lived in urban areas and this study was underpowered to analyze rural food environments. BCGP participants were volunteers, which introduces the risk of participation bias and limits generalizability. BCGP participants tend to be higher income, more educated, older, and generally healthier (e.g. lower prevalence of smoking) than the general population in BC (38,39). However, it is notable, that even in this typically higher SES study population, participants who lived in neighbourhood with greater density of fast-food restaurants were less likely to meet FV recommendations. Potential disconnects in the timing of data should also be acknowledged. Can-FED measures were derived from 2018 Business Registry data (20), CANUE variables were derived from the 2016 Census, while BCGP participants were recruited between 2009-2016 (19). However, since food and neighborhood environments evolve slowly, any impact on associations were likely negligible. Consumption of FV was self-reported which is prone to underreporting (40). Additionally, BCGP did not collect dietary information beyond FV consumption which limits the ability to assess associations between the food environment and diet quality more broadly which may be particularly insightful in the context of fast-food restaurants and convenience stores.

## Conclusion

Our findings suggest that food deserts are prevalent across urban areas in BC. Conversely, areas with high density of fast-food outlets and convenience stores are rare. Living in neighbourhoods with a higher density of fast-food outlets was associated with lower FV consumption in this population. Further studies are needed to understand the null findings, particularly, whether the higher SES population in BCGP resulted in smaller effect estimates for the food environment on dietary intake. Studies should also incorporate additional factors that shape dietary intake such as food affordability and consider the influence of the food environment where people work and spend leisure time, in addition to where they live.

## Supplementary Materials

Table S1: Descriptive statistics of participant characteristics by category of FV consumption. Table S2: FV consumption among classes of food environment measures.

## Author Contributions

Conceptualization, BZ, RAM; methodology, BZ, RAM, JS; formal analysis, BZ; investigation, BZ; resources, RAM, PB; writing—original draft preparation, BZ, RAM.; writing—review and editing, TRC, RM, PB, JS.; supervision, RAM, JS, PB, TRC, RM. All authors have read and agreed to the published version of the manuscript.

## Data Availability Statement

Software code is available from researchers upon reasonable request. Code used to derive variables is returned to the BCGP as per data access requirements. The datasets analyzed in this study were made available by the BCGP and are available on reasonable request and with approval from the BCGP.

## Acknowledgments

A version of this work was published as part of Bill Zhao’s Masters thesis. https://dx.doi.org/10.14288/1.0447286. The BCGP is only possible with the commitment of its research participants, its staff, and its funders. The data used in this research were made available by the BC Generations Project.

